# The Failure Index as marker of cochlear health in MED-EL CI users: Anatomy, demographics, and speech correlations

**DOI:** 10.64898/2025.12.02.25341153

**Authors:** Wiebke Konerding, Cornelia Batsoulis, Peter Baumhoff, Heval Benav, Lutz Gärtner, Annette Günther, Onhintz de Olano Dieterich, Daniel Schurzig, Stefan Strahl, Jochen Tillein, Sarah Vormelcher, Andreas Büchner, Carolyn Garnham, Andrej Kral

**Affiliations:** Department of Experimental Otology, Hannover Medical School, Hannover, Germany; MED-EL Research Center, MED-EL Medical Electronics GmbH, Hannover, Germany; Research & Development, MED-EL Medical Electronics GmbH, Innsbruck, Austria; Department of Otolaryngology, Hannover Medical School, Hannover, Germany; Clinics of Otolaryngology, Head and Neck Surgery, J.W. Goethe University, Frankfurt, Germany

**Author notes:** Contributed equally (Andreas Büchner, Carolyn Garnham, Andrej Kral).

**Keywords:** neural health marker, electrically-evoked compound action potential, objective measure, cochlear implant

## Abstract

Cochlear implants (CIs) enable hearing with the deafened ear, via direct, electrical stimulation of the spiral ganglion neurons (SGN). Thus, the outcome depends on the number and excitability of the SGNs. We recently established the electrically-evoked compound action potential (eCAP)-derived Failure Index (FI) as cochlear-health marker in the animal model. The FI informs about the presence, site, and size of a SGN lesion.

Here, we translated the FI to clinical recordings of MED-EL CI users. For the retrospective study, we selected patient data from the database of the German Hearing Center Hannover recorded 2017 to 2024. We included 199 post-lingually and 79 pre-lingually deafened ears.

Averaged FIs over all contacts of a CI were stable within the analysis period (3^rd^ month to 1^st^ year postoperatively). The FI increased with age and was elevated for etiologies associated with higher SGN loss. Utilizing 3D information from cone beam-computed tomography scans, we confirmed that the FI was independent of distance (0.1-2.5 mm) to the modiolus. The FI showed individual patterns along the array with maxima usually at basal contacts, corresponding to elevated SGN loss at high frequencies. In a selected group of post-lingually deaf ears, we confirmed the correlation of the FI with speech perception in quiet and in noise (n=28, r^2^=0.12-0.55).

Thus, we propose the FI as promising clinical tool to identify CI-implanted ears with reduced neural health and contacts close to areas of SGN loss. Thereby, it can serve to guide speech-processor fitting to optimize CI outcomes.

## Introduction

Cochlear implants (CI) enable speech understanding in deafened subjects, both post-lingually implanted adults (e.g., Sargsyan et al., 2021) and early implanted pre-lingually deaf children (e.g., Illg et al., 2024). However, the outcomes in speech understanding vary substantially (Boisvert et al., 2020; Illg et al., 2024) and especially understanding speech in background noise remains a challenge for many CI users (for review see Henry et al., 2021). Predictive factors are scarce and explain only part of the outcome variability (Demyanchuk et al., 2025; Günther et al., 2025; Zhao et al., 2020). Besides cognitive-linguistic factors (e.g., Zhan et al., 2020) the factors repeatedly reported to influence speech understanding are age, and duration of deafness (e.g., Heutink et al., 2021; O’Donoghue et al., 2000).

One important peripheral factor is neural health, which is the survival and function of the target neurons of the CI. However, as there is no possibility to assess spiral ganglion neuron (SGN) counts *in vivo*, a reliable electrophysiologic marker is needed (Zhan et al., 2021). A neural health marker would allow to predict the best possible outcomes with a CI, and could be used to tailor speech-processing programming to the individual requirements. A recent review among audiologists reported that although a majority believes that silencing contacts close to areas of reduced neural health would improve speech understanding, this adjustment is rarely performed, as guidelines for selecting contacts are missing (Sander et al., 2023). Thus, a current focus in CI research is to find a marker to guide tailored speech-processor programming (for review see Lien et al., 2025). A common approach is to assess characteristics of the electrically-evoked compound action potential (eCAP) that can be recorded non-invasively via the CI (for review see Schvartz-Leyzac et al., 2023). In humans, one verification of a neural health marker is a significant correlation with speech understanding scores. So far, the attempts led to mixed results, with eCAP threshold in monopolar stimulation mode usually failing to reach significant correlation with speech outcomes (but see focused threshold, e.g., Arjmandi et al., 2022; Peng et al., 2025) and other proposed markers usually explaining less than 30% of the variance in speech understanding (for review see Dawson et al., 2025; Schvartz-Leyzac et al., 2023, 2025). For different electrophysiological markers, the correlation was reported to be stronger for speech perception in noise than in quiet listening (Schvartz-Leyzac & Pfingst, 2018; Skidmore et al., 2022, 2023).

Several studies reported that high variability of threshold levels across electrodes can negatively impact speech recognition (DeVries et al., 2016; Pfingst et al., 2004; Zhou & Pfingst, 2014). Thus, a neural health marker needs to be sensitive to local differences in neural health in order to reliably reflect outcome variances. In a novel approach, we established a neural health marker, the Failure Index (FI), in the animal model (Konerding et al., 2025). The FI is calculated as the input/output ratio at maximal activation, indicating the failure to transform electrical current into neural signals (i.e., synchronous activation). The rationale for calculating the quotient between amplitude and current, is its normalizing effect for current spread, thus generating a comparable matrix between different contacts and ears. In the guinea pig, the FI separated lesioned-from non-lesioned ears at the overall median (i.e., grand median, FI_GM_), allowing an identification of reduced neural health at the individual level. Furthermore, the maximum of the FI along a CI indicated the contact closest to the lesion in 80% of cases (i.e., except for small lesion sizes).

In the guinea pig, the FI was a more reliable predictor of reduced neural health than other characteristics of the eCAP amplitude-growth function (AGF), such as threshold, or maximal amplitude. We hypothesise that the (partial) normalization for current spread is reducing the influence of distance to excitable structures. In contrast, both threshold and amplitude, were found to be dependent on electrode-to-modiolar distance (EMD; Degen et al., 2020). In this retrospective study, we translated the FI to human CI users (all implanted with MED-EL device) to test this hypothesis and to assess dependency on demographic factors linked to SGN loss. We assessed the translational potential of the FI according to 5 different criteria: 1) Stability over time, 2) group differences (i.e., age and etiology), 3) correlation with speech perception outcomes, 4) individual profiles along the electrode array, and 5) independency from anatomical position (i.e., EMD).

## Material and Methods

### Subjects

For this retrospective cohort study, we selected 278 CI implanted ears (MED-EL GmbH, Innsbruck, Austria) of 253 CI users (median age: 54 years; 140 females; classification as per self-identification; see patient demographics in Appendix 1). The age was defined as age at implantation as all measurements were performed within the same time-period from 3^rd^ month (M3) to 1^st^ year (Y1) following implantation.

All included CI users received a flexible electrode array (MED-EL: FLEX series) selected to match the individual cochlear anatomy. In case of simultaneous binaural implantation, both ears were included and analysed individually. Exclusion criteria were second ears (n=91), atypical implant types (i.e., n=6 short arrays of FLEX16 and FLEX20, and n=7 uses of stiff electrode arrays), stem cell applications (n=6), and reimplanted ears (n=14). With exception of the assessment of anatomical relations (see ‘Correlation with electrode-to-modiolus distance’), ears with partial insertion of the electrode array (n=49) were excluded. Thus, a total of 197 post-lingually and 79 pre-lingually deafened ears were included in the final analysis of the full data-set. To assess the correlation with outcome measures, we chose additional exclusion criteria to reduce variability due to differences in language experience and number of contacts included in the analysis (for details on selection of the reduced data-set see ‘Correlation with speech scores’). Due to differences in the underlying causes of deafness that impacted the outcome measure as well as differences in speech understanding we differentiated between pre- and post-lingually deafened ears, where necessary (details below).

The CI surgeries were performed at the Hannover Medical School (MHH) from August 16^th^, 2016 to February 2^nd^, 2024 and the post-implantation data was collected during clinical visits at the German Hearing Center Hannover (i.e., tertiary care centre, specialized in the treatment of hearing loss). Objective clinical and anatomical data were integrated into the CI database (for details see, Günther et al., 2025).

As there was no significant difference in average FI between males and females (Mann Whitney: U=8603, p=0.1699), left and right ears (Mann Whitney: U=9188, p=0.5491), and whether or not the CI was used with an additional acoustic component (Mann Whitney: U=2212, p=0.9839), we combined all ears for further analyses, selecting sub-groups of the dataset according to the particular analysis made (detailed below).

### ECAP recordings

The response of the auditory nerve to CI stimulation was assessed in terms of the eCAP N1-P1 amplitude, using the automatized, continuous eCAP measurement function ‘AutoART’ (Strahl et al., 2018) of the MAESTRO fitting software (Version 7 or higher, MED-EL, Innsbruck, Austria). In contrast to the animal model, where recording was performed via an extra-cochlear electrode at the round window, referenced to a subcutaneous electrode behind the ear (Konerding et al., 2025) the eCAP in MED-EL CI users was recorded via a CI contact referenced to an external ground contact located on the body of the implant package, outside the skull. We always chose the CI contact(s) adjacent to the stimulation contact for recording.

Stimulation was delivered to each of the 12 contacts, separately, with biphasic single pulses of opposite leading-phase polarity for artefact reduction (for details see Konerding et al., 2022, 2025). Whereby, slightly different pulses were used with phase-durations ranging from 40s to 60µs and interphase-gaps of either 2.1µs or 10µs. The amplitude of the electric stimulation in proprietary charge units (qu∼nC) was steadily increased either until the subject indicated that the sound was too loud, or until a pre-defined safe-charge limit of each electrode contact (max 72nC) was reached.

The resulting amplitude growth function (AGF) was then fitted with a sigmoidal function to assess the parameters used to derive the FI (Fig. 1A). Since there were eCAP AGFs from both adjacent recording contacts for most stimulating contacts, we calculated the mean parameters for threshold, amplitude and FI. We restricted the analysis to those AGFs for which the automated procedure of the recording software detected a threshold. This criterion led to a drop-out of 6% of the 278 ears and 27% of individual contacts.

**Fig. 1.**
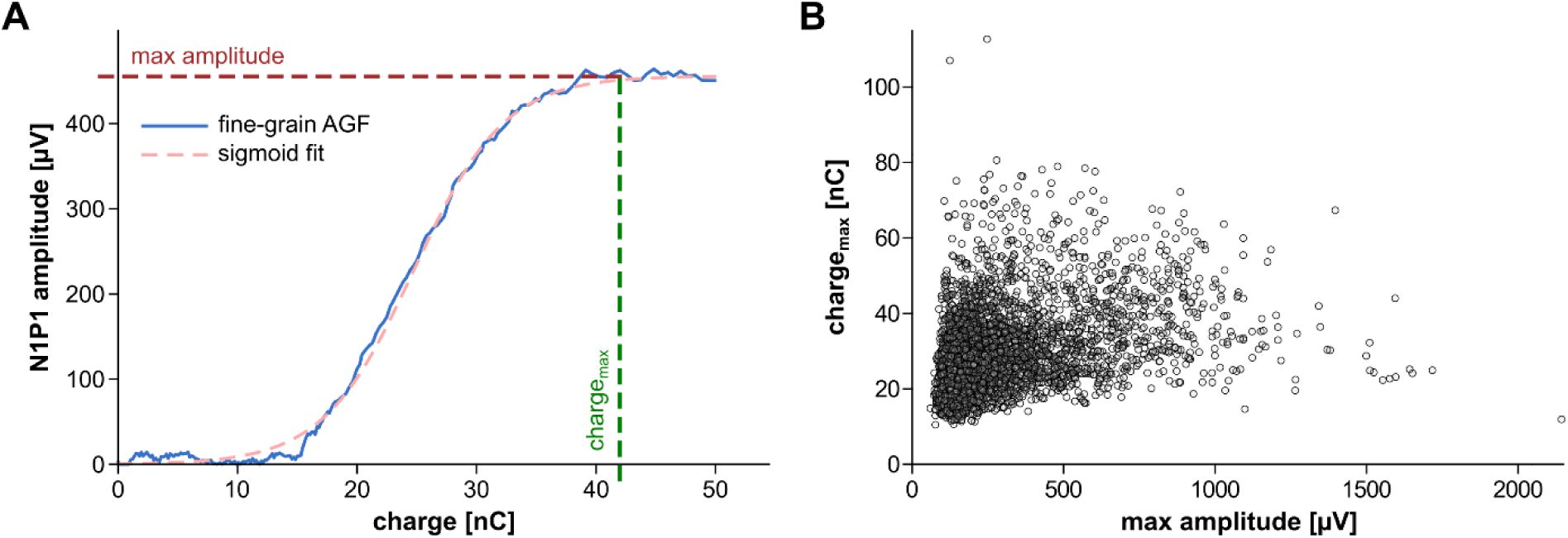
A) Illustration of the procedure to derive the parameters for FI calculation from the AutoART fine-grain amplitude-growth function (AGF). Given is a representative example (blue) and the resulting sigmoidal function fitted to the AGF (red curve). The FI (here: 0.09 nC/µV) is calculated from the maximal N1P1 amplitude (red line) and the minimal charge level needed to reach saturation (charge_max;_ green line). B) The depiction of all 4082 corresponding maximal amplitudes and charge_max_-levels included in the study, highlights the broad range of values in each of the two dimensions used to calculate the FI.

### Failure Index

The FI was calculated based on the AGF characteristics maximal amplitude (i.e., fitted upper asymptote) and the minimum charge needed to reach saturation (i.e., charge_max_ at 99% of the upper asymptote; formula 1; Fig. 1). Differently from the animal study (Konerding et al., 2025), we did not use current, but charge (in nC) to incorporate different phase durations.

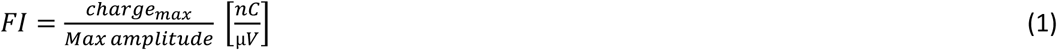

### Correlation with speech scores

For this comparison, we chose a reduced data set, including only post-lingually deaf patients who used electrical stimulation only. Patients with combined electric-acoustic hearing, either using an amplified acoustic or a normal/natural hearing in low-frequencies were excluded. We excluded non-native speakers (n=12) and patients suffering from comorbidities that could affect speech understanding (n=5; i.e., tinnitus, mental retardation and ‘difficulty finding words’). The resulting sample sizes differed based on speech-perception tasks (since not all tests were carried out in all subjects). The respective numbers are given in the results section.

The most commonly performed test in our sample was the Freiburger monosyllabic word test (Hahlbrock, 1953; Hochmair-Desoyer et al., 1997) performed in quiet. Additionally, we included the German Hochmair-Schulz-Moser (HSM) sentence test both in quiet and in noise. The HSM test consists of 30 lists of 20 everyday sentences, each with 106 words (Hochmair-Desoyer et al., 1997). For the HSM test in noise, a noise with a speech-shaped spectrum was added at −10 dB signal-to-noise ratio. The acoustic stimuli were presented from a loudspeaker and perceived by the subjects via their everyday speech processor strategies. In case of single-sided deafness the second ear was blocked or masked during the task. The perceptual outcomes were scored as percentage correct words. Due to the demands resulting from fast progression of sentences within the test, subjects were sometimes observed to withdraw from the task. Thus, we decided to include only non-zero HSM scores (i.e., exclusion of floor effects).

Due to the collinearity between the different CI-contacts (variance inflation factor, VIF >5), it was not possible to combine all FI values in one linear model to predict speech understanding scores. Thus, we calculated (analogous to the guinea pig study; Konerding et al., 2025) the maximal (worst) FI for each ear and correlated it with the maximal (best) speech scores during the same investigation period (M3-Y1). Ideally, the maximal FI would be calculated over all 12 CI contacts. As this restriction would lead to further reduction in sample size, we chose those ears for which the same 10 contacts delivered an FI (i.e., contact #1 through #10). We confirmed that the resulting linear equation for the correlation with highest coefficient of determination was similar to the one using the subset of ears with all n=12 contacts included (n=20). To put the resulting speech correlations into perspective, we also correlated the speech scores with eCAP threshold and maximal amplitude (i.e., the denominator in the FI calculation, formula 1).

### Correlation with electrode-to-modiolus distance (EMD)

As an approximation of the distance of the CI contacts to the excitable structures we calculated the EMD from the centre of the electrode contact to the modiolar wall (Garcia & Carlyon, 2025; Salcher et al., 2021). The 3D reconstructions of the cochleae were performed based on cone beam-computed tomography datasets (Xoran XCAT or the Morita ENT devices) recorded during the clinical routine at the Hannover Medical School: Initially, segmentations of both the lateral cochlear wall within preoperative imaging (Meng et al., 2016; Schurzig, Timm, Lexow et al., 2018; Würfel et al., 2014) and of the CI contact positions in postoperative imaging were performed, as previously described (Salcher et al., 2021; Timm et al., 2018), using the software tool OsiriXMD (version 2.5.1 64bit, Pixmeo SARL, Switzerland). These segmentations were registered as described in Schurzig et al. (2018), and the shape of both the inserted electrode array as well as the scala tympani were reconstructed using the same methodology as in the recently proposed virtual implantation (Schurzig et al., 2023), yielding a geometrical representation from which the EMD of the individual contacts was extracted (Fig. 2).

**Fig. 2.**
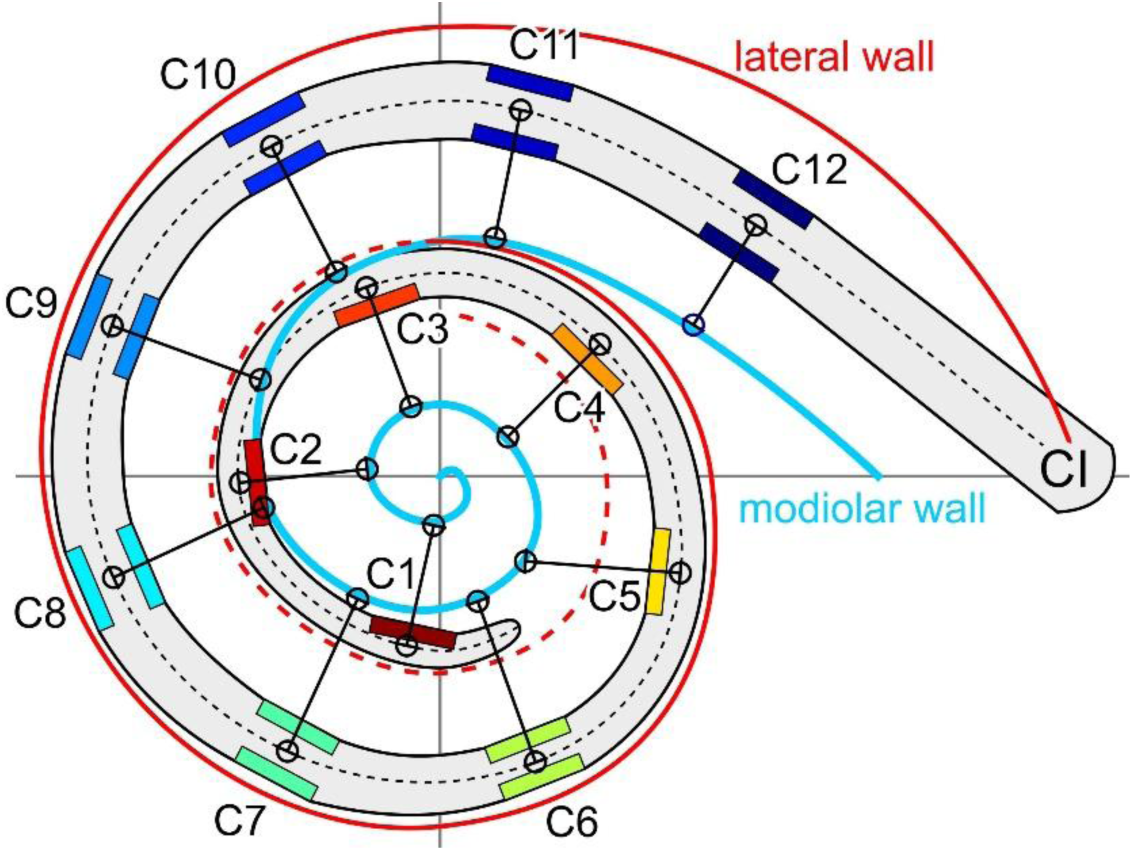
Assessment of electrode-to-modiolar wall distance (EMD) from 3D reconstructions of the cochlea. Indicated is the reconstruction of the scala tympani, overlain by the reconstructed CI array from basal contact C1 to apical contact C12. The distance (black, connecting line) was measured from the midpoint of the array at the position of each contact to the edge of the scala tympani. The modiolar wall (blue) was reconstructed based on scala width from the lateral wall (red; dashed line indicates the progression of the scala wall covered by more apical turns).

As with the correlation to perceptual outcomes, we assessed the dependency on EMD not only for the FI, but also for the threshold and the maximal amplitude.

### Statistics

Statistical tests were performed using GraphPadPrism 5 (GraphPad Software, Inc., Boston, MA, USA). For group analyses, we performed t-tests (e.g., sex), 1-way ANOVA (e.g., cause of deafness), or 2-way ANOVA (e.g., contact and recording time). To correct for multiple testing, we chose Bonferroni correction in the post-test comparisons. Wherever applicable, we used non-parametric tests (e.g., indicated by non-significant Kolmogorov-Smirnov test). In order to assess linear relations (correlations with speech scores), we used Pearson correlations and additionally report linear regression coefficients for significant correlations (e.g., speech). For non-linear correlations (e.g., age) we performed Spearman correlations and indicate regression curves with correlation coefficients for significant correlations.

## Results

The median FI over all 199 post-lingually deaf ears (median over all contacts of the CI, averaged for the study period) was 0.147 nC/µV and the median over all 79 pre-lingually deaf ears was 0.136 nC/µV. As there was no significant difference between pre- and post-lingually deaf ears in average FI (Mann-Whitney: U=7346; p=0,3952), we combined them for dichotomization of the data: To distinguish at the group level between presumed healthy auditory nerves and cochleae with presumed reduced neural health, we used the grand median FI_GM_=0.143 nC/µV (i.e., median over all ears; DeCoster et al., 2011) and defined those with mean FI<FI_GM_ as low FI (i.e., presumed health ears) and those with mean FI≥FI_GM_ as high FI (i.e., ears with presumed SGN loss).

### Stability over time

To take into account the individual differences between contacts, we assessed the influence of recording time (i.e., 3, 6 and 12 months post-implantation) on FI with a 2-way ANOVA, combining contacts and time. This analysis was performed for a subset of 70 ears, which were tested at all 3 time points (Fig. 3A). Whereas there was an overall significant difference between contacts (details below), there was no significant difference based on recording time from M3 till Y1 post-surgery, and also no interaction between recording time and contact (Contact: F(1476,11)=16.08, p<0.0001; Time: F(1476,2)=0.0648, p=0.9373; Interaction: F(1476,22)=0.1972, p=1.000; Fig. 3). Due to the stability over time, we could average the FI over the analysis period (M3-Y1), which increased overall statistical power, as not all ears were tested at every time point.

**Fig. 3.**
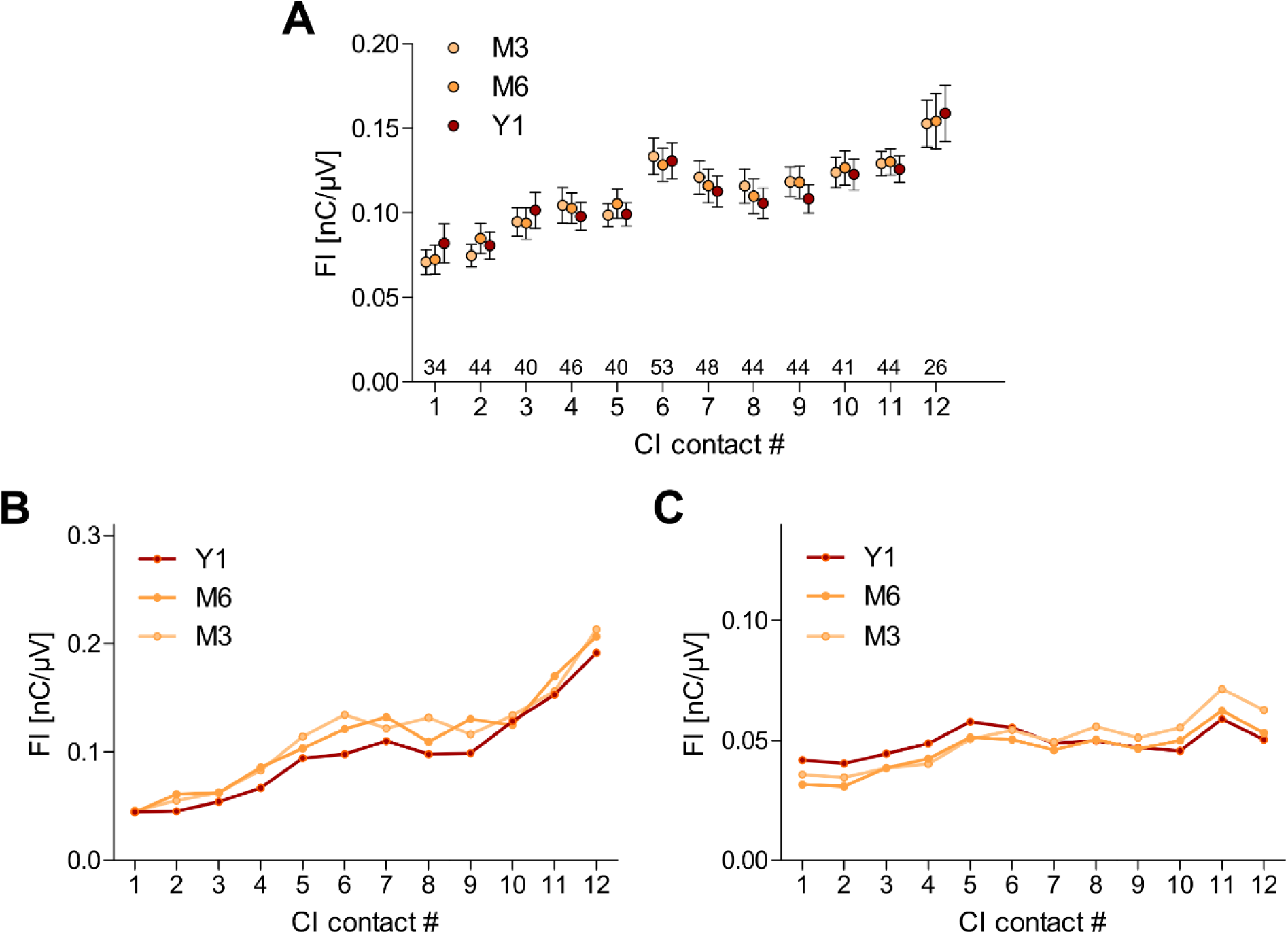
A) Stability of FI-values over time from recording at month three (M3), over month 6 (M6) to year one (Y1) post-surgery. Given are mean-values with the standard error of the mean (SEM). Number of ears per contact are given above the abscissa. B&C) Two representative ears (selected from A) with low mean FI (B: 0.05 nC/µV, C: 0.10-0.11 nC/µV) with distinct, but stable patterns along the array. Note the difference in axis scaling.

### Age-Dependency

We analysed the correlation of the mean FI per ear with age at implantation. As the causes of deafness differed between pre- and post-lingually deaf ears (details below), we restricted the analysis to cases with idiopathic etiologies. We further excluded ears (n=16) which had received treatment (i.e., short-or long-term local steroid application) that may alter age-typical changes in neural health. There was a significant increase of FI with age for combined pre- and post-lingually deaf ears (Pearson r=0.268, p=0.0009, n=150). The regression yielded higher coefficients of determination for non-linear (i.e., sigmoid: r^2^=0.132) than linear regression (r^2^=0.072), indicating a rapid rather than gradual change in FI with age above 60 years of age. When binning the data per age group (bin-size: 20 years), we revealed a significant increase in FI for the group 60-79 years relative to the youngest age group (i.e., <20y; 1-way ANOVA: F(4,145)=4.478, p=0.0019 with Bonferroni corrected post-test; Fig. 4). Thereby, the FI was crossing the line from presumed healthy auditory neurons (<60 year: average FI <0.143) to presumed reduction in neural health after 60 years of age.

**Fig. 4.**
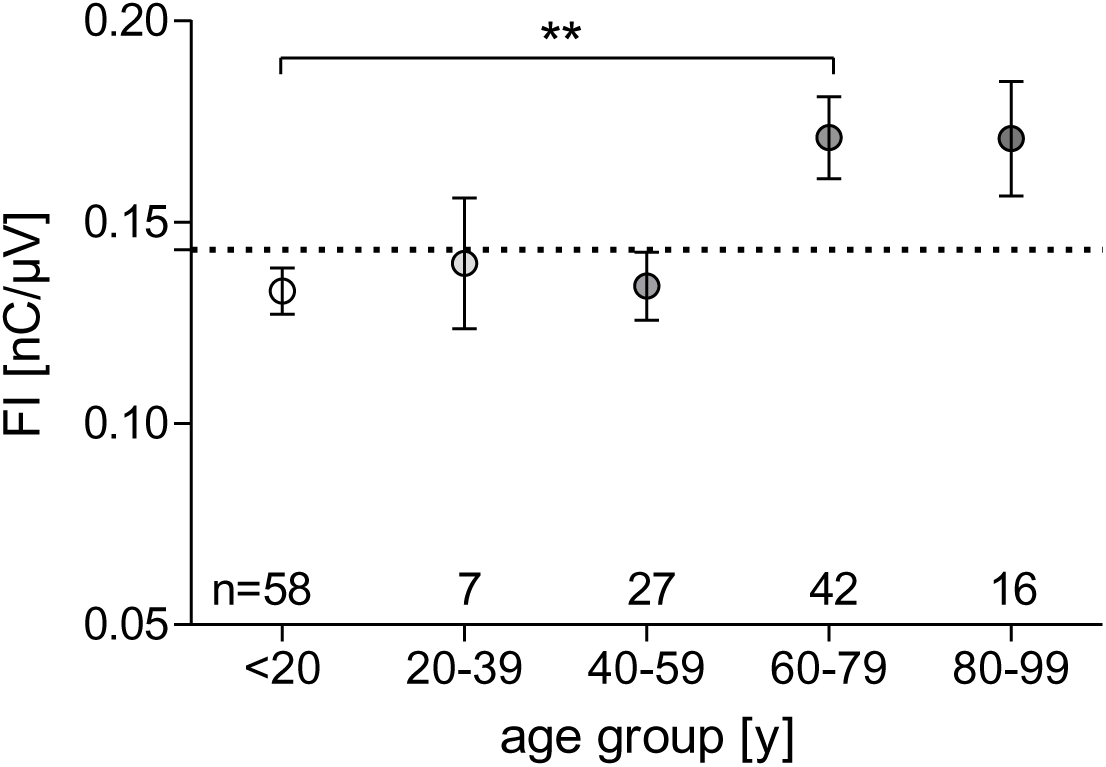
Increase of average FI (mean over M3-Y1) with age for combined pre- and post-lingually deaf ears. Given are means (symbols) and SEM (whiskers) for each age group (bin-size: 20 years). Sample sizes per group are indicated above the abscissa. The dashed line indicates FI_GM_=0.143 nC/µV used for dichotomization of the data into presumed healthy auditory nerves (below) and cochleae with reduced neural health (above the line).

### Influences by etiology

For most ears, the cause of deafness was unknown/ idiopathic (109 post-lingual, 57 pre-lingual) and the majority of causes were unique to either post- or pre-lingual cases. The most common cause was sudden hearing loss (n=45, only post-lingual), followed by genetic causes (6 post-lingual, 8 pre-lingual), Menière’s disease (n=12, only post-lingual), cochlear nerve hypoplasia (n=7, only pre-lingual), and traumatic incidences (n=6, only post-lingual). Due to these distinctions and the described changes in FI with age, we assessed the influence of etiology on FI for post- and pre-lingually deaf ears, separately. We excluded ears which had received steroid treatment, as we did for the correlation with age.

In the post-lingually deaf group there was a significant difference in FI based on etiology (1-way ANOVA: F=3.720, p=0.0164, df=3; sample sizes see Fig. 5) with deafness of genetic origin leading to significantly increased FI compared to sudden hearing loss (sudden HL, Fig. 5A). In the pre-lingually deaf group, the cochleae with hypoplastic auditory nerves had significantly elevated FI as compared to cochlea with genetic cause of deafness (Mann Whitney: U=4.000, p=0.0065, Fig 5B). Furthermore, the hypoplasia group was significantly elevated above the FI_GM_ (Wilcoxon test vs FI_GM_: W=26, p=0.0313), with the lowest value (FI=0.141nC/µV) being close to line between supposed healthy and affected ears (FI_GM_=0.143). The difference in FI between the post- and the pre-lingual ears with genetic causes (Fig. 5 A&B – red) is likely attributed to older age in those with post-lingual (35 to 76 years) as compared to pre-lingual onset of deafness (1 to 22 years). Concordantly, there was a significant increase in FI with increasing age at implantation for these 13 ears with genetic cause of deafness (Pearson r=0.658, p=0.0146; linear regression: r^2^=0.433).

**Fig. 5.**
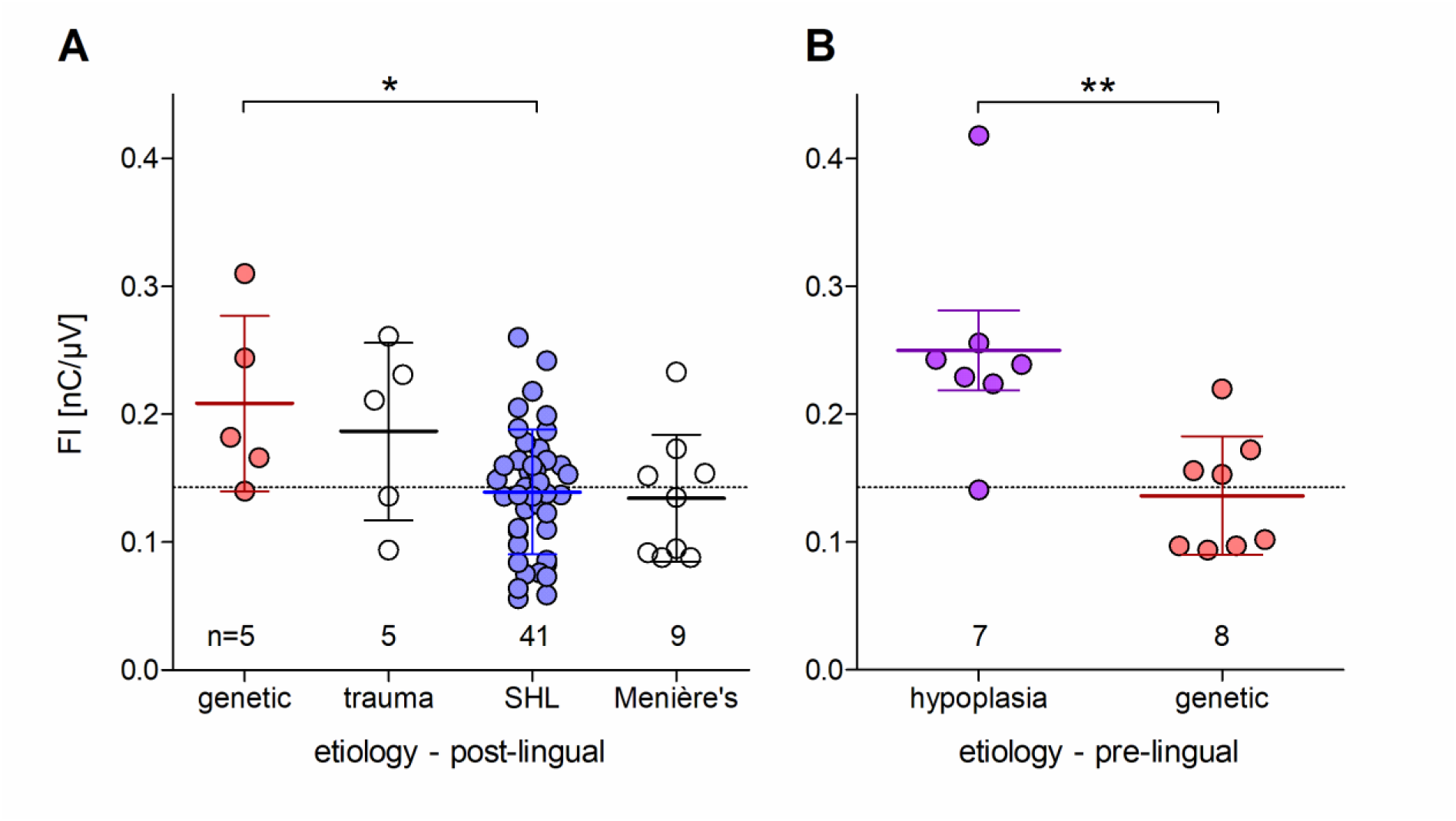
Differences in FI (mean over M3-Y1) based on most-common etiologies within the study’s database for A) post-lingually and B) pre-lingually deafened ears. Given are group means (lines) and SD (whiskers), overlaying single cases (symbols). Sample sizes per group are indicated above the abscissa. The dashed line indicates FI_GM_=0.143 nC/µV used for dichotomization of the data into presumed healthy auditory nerves (below) and cochlea with reduced neural health (above the line). SHL – sudden hearing loss

### Dependency on stimulation contact

Based on the findings in the animal model (Konerding et al., 2025), we hypothesised that FI maxima along the array for ears with mean FI above FI_GM_ (i.e., mean FI_M3-Y1_>0.143) indicate regions with reduced neural health close to the respective stimulation contact. Therefore, we selected ears with high average FI for which at least 11 contacts were available. The maxima of these 36 ears showed a wide variability of positions from contact #1 to contact #12 with most cases peaking at basal contact #12 (n=10), followed by contacts #9 and #11 (n=6, each; Fig 6A). In two representative cases (Fig. 6 B&C), for which data was available at both M3 and Y1, the overall stability of the peaks and troughs in FI pattern along the array was confirmed, albeit with increasing average FI over time. This increase from M3 to Y1 post-surgery was specific to these cases of assumed low neural health (i.e., high mean FI) and opposed the general stability of the FI over time (Fig. 2).

**Fig. 6.**
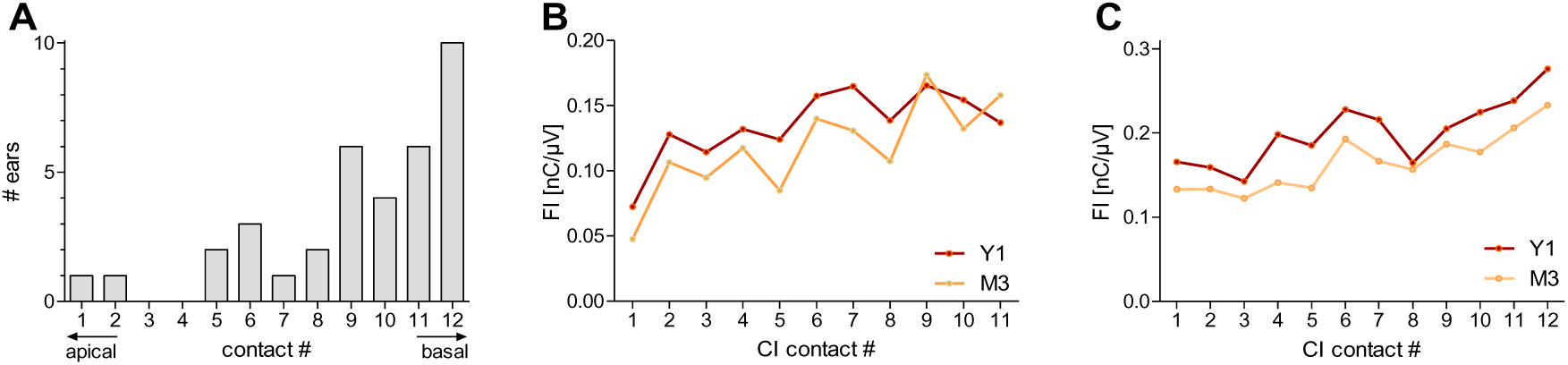
A) Histogram of the position of FI-maxima along the array from contact number one (most apical) to contact number 12 (most basal) for ears with overall high FI (i.e., mean FI> 0.143; N=36). B&C) Two representative ears with high mean FI at Y1 (A: 0.14, B: 0.20) show distinct and reproducible FI-pattern with multiple peaks and troughs along the array at three months (M3) and one year (Y1) post-surgery. Note the difference in scaling for B&C).

### Speech correlation

Inferring from the findings from the animal model (Konerding et al., 2025), we expected a correlation of maximal FI-values with severity of hearing impairment, assessed here in terms of speech understanding scores. For both speech understanding and FI, we used the maximum (highest score) over all measurements from M3 to Y1.

We restricted the analysis to a reduced data set of post-lingually deaf ears for which the same 10 contacts were available for calculating the maximal FI. All tested speech outcomes significantly decreased with increasing FI: monosyllabic word scores (Pearson r=-0.345, p=0.0273, N=41; Fig. 7A), HSM sentence scores in quiet (r=-0.532, p=0.0043, N=27; Fig. 7B), and HSM sentence scores in noise (r=-0.738, p<0.0001, N=28; Fig. 7C). We confirmed that the regression line derived for this reduced data set, was similar to the one using an even smaller subset of ears (n=20) for which all 12 contacts were available (HSM score in noise: n=10 contacts: y = −220*x + 100; n=12 contacts: y = −203*x + 100; Fig. 7C).

**Fig. 7.**
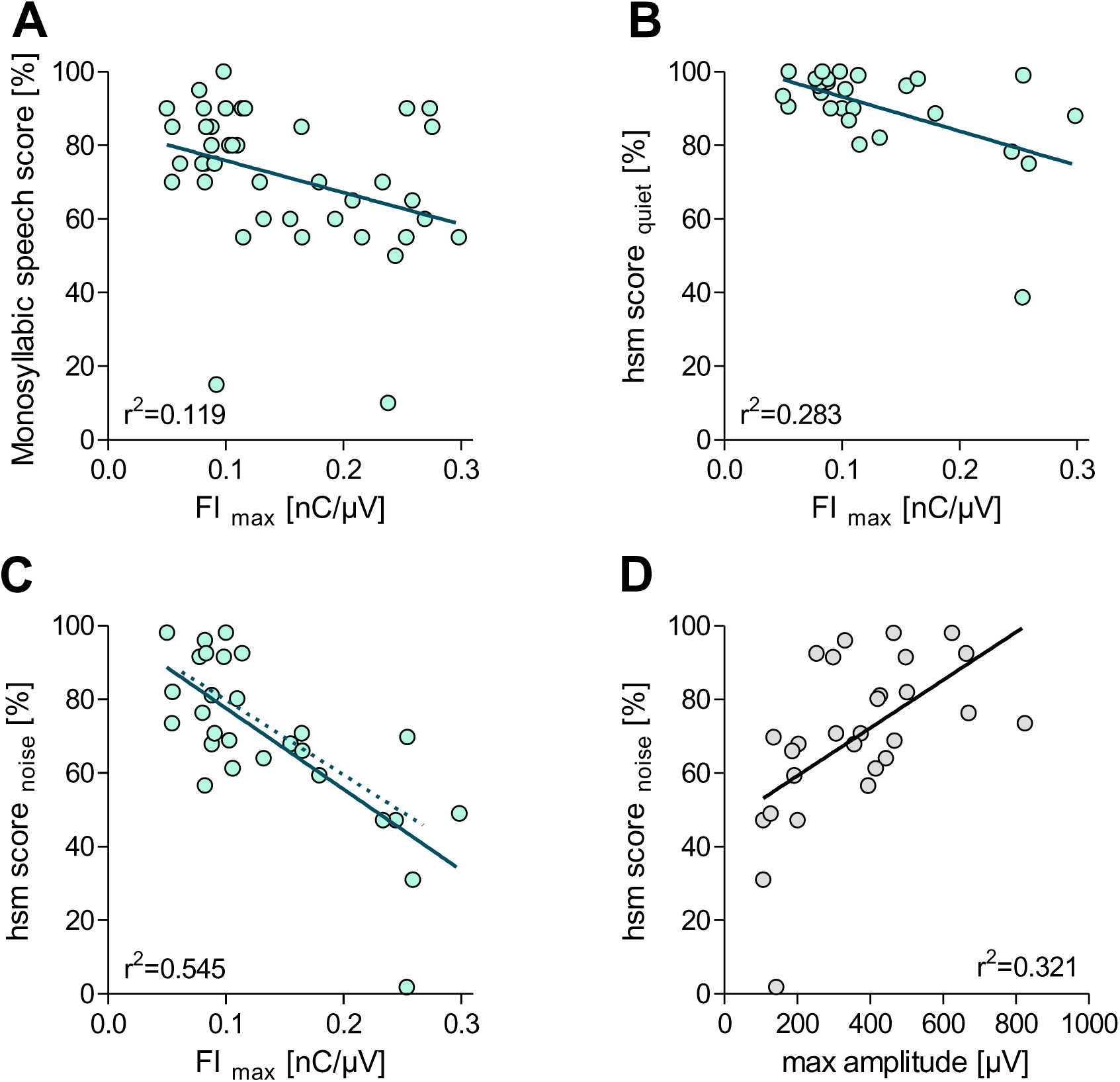
Correlation of eCAP measures with speech test outcomes for ears for which the same n=10 contacts were available to calculate the extreme values along an array (i.e., maximal speech score, maximal FI and minimal value of the maximal eCAP amplitude; all for M3-Y1). The FI correlated significantly with A) monosyllabic speech scores, B) HSM scores in quiet, and C) HSM scores in noise. D) The maximal eCAP amplitude significantly correlated with HSM scores in noise. A-D) Given are the linear regression line (solid line) and the respective coefficient of determination (r^2^). C) The additional linear regression line (dashed line) indicates the dependency of FI on HSM score in noise when the maximal FI was calculated based on a subset of ears for which all 12 contacts were available.

We further assessed the potential correlation of eCAP thresholds and maximal eCAP amplitude (i.e., minimum over 10 contacts) with speech outcomes. There was no dependency of speech outcome on threshold, for any of the three tests (monosyllabic: Pearson r=0.152, p=0.3425; HSM quiet: r=0.129, p=0.5217; HSM noise: r=-0.057, p=0.7751). We confirmed the expected correlation with maximal amplitude for the word test (monosyllabic: Pearson r=0.405, p=0.0087) and sentence test in noise (HSM noise: r=-0.057, p=0.7751), but not sentence test in quiet (HSM quiet: r= 0.154, p=0.4426). Furthermore, the variance in maximal amplitude explained at most 32% of the variance in speech outcome (r^2^=0.321; Fig. 7D) whereas the variance in FI explained more than half of it (r^2^=0.545; Fig 7C) in the study cohort.

### Independency of FI on EMD

We assessed the dependency of eCAP parameters on distance to excitable structures by correlating the parameters (as a group) with the EMD. For this, we chose all data available in our data base, including also partially inserted CIs (N=1907 contacts; Fig. 8A & B: grey symbols) and repeated the analysis with the subset of ears (n=28) that was used for speech-in-noise correlation (i.e., all full insertions; N=251 contacts; Fig. 8B: pink symbols). The EMD did not change steadily with insertion angle, but reached a maximum at around 300° (i.e., disregarding the contacts situated close to the round window at ∼0°; Fig. 8A). Due to differences in morphology and insertion depth, the contact number (#1-12) did not predict the insertion angle (Fig 8B). Thus, each contact was included independently in the correlation analysis (i.e., for the group). Because the correlation with speech-test outcomes was based on a reduced data set of ears that all had full insertions, we repeated the analysis restricted to these ears.

**Fig. 8.**
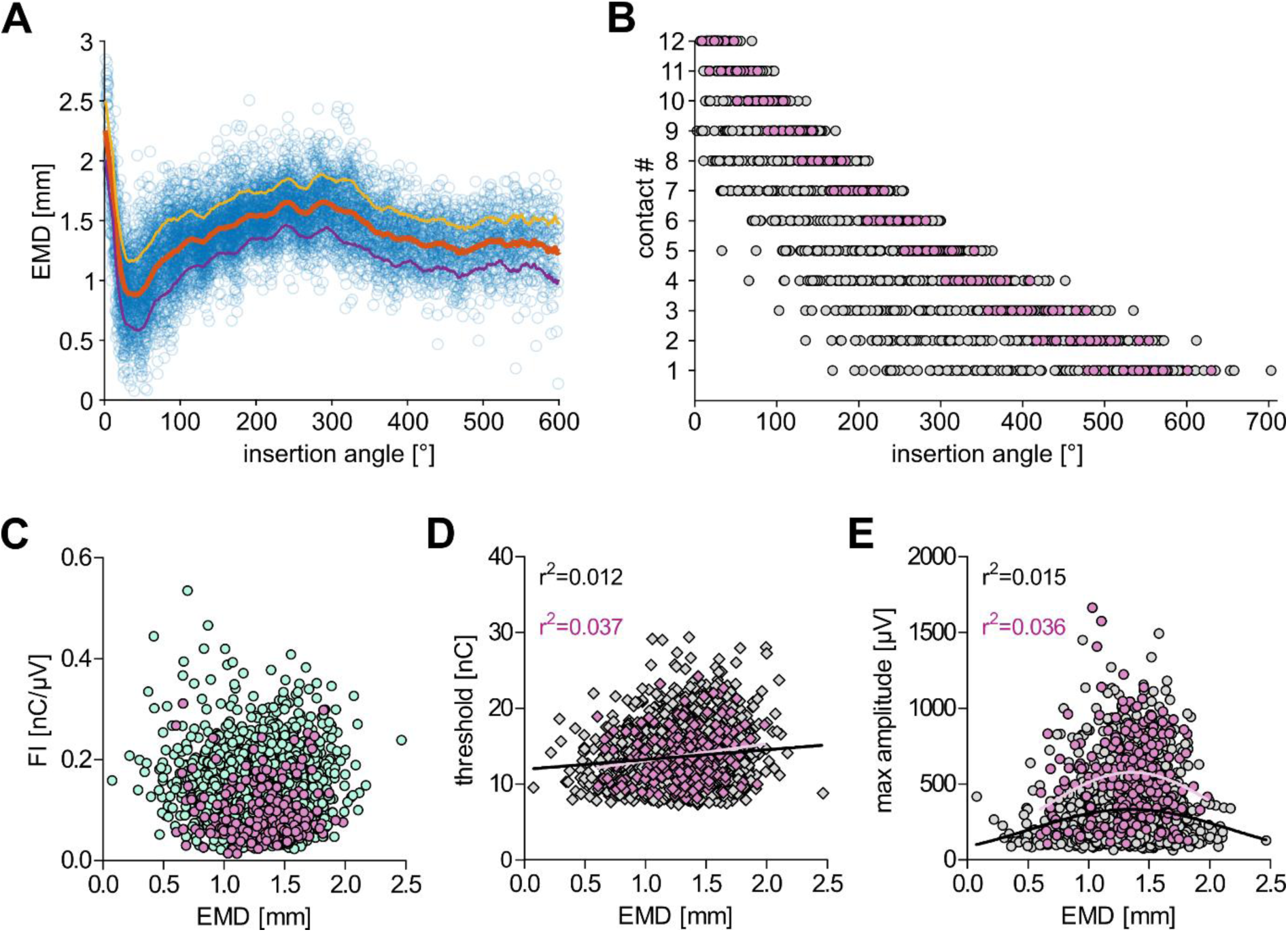
A) The electrode-to-modiolar wall distance (EMD) changed non-monotonically with insertion angle. B) The contacts in the full data set (grey symbols) covered a wide range of insertion angles per contact number. This spread was substantially reduced for a subset of data with full insertions (pink symbols; selection criteria as per speech correlation; Fig. 7C). C-E) The dependency of the different eCAP parameters on EMD differed according to our hypotheses: C) No correlation with FI, D) correlation with eCAP threshold and E) with maximal eCAP amplitude. D & E) The linear (threshold) and non-linear regressions (max amplitude) and the resulting coefficients of determination are given for both the full (black) and the reduced data set (pink).

The FI did not change systematically with EMD, both for the full data set, including partial insertions (Spearman r=-0.025, p=0.2666, Fig 8C) and the reduced data set (Spearman r=-0.026, p=0.687). The eCAP threshold was significantly affected by distance, with a linear increase in threshold with increasing distance from the modiolar wall both for the full data set (Spearman r=0.088, p=0.0001; Fig 8D) and the reduced data set (Spearman r=0.198, p=0.0018). As expected, the maximal eCAP amplitude showed an intermediate dependency on EMD, which resulted in a significant correlation in the full data set, with highest amplitudes for intermediate EMDs (Spearman r=0.055, p=0.0163; gaussian fit; Fig 8E) and a lack of significant correlation in the reduced data set (Spearman r=0.110, p=0.0860).

## Discussion

We assessed and confirmed the translational potential of the Failure Index (FI) according to 5 different criteria: 1) Stability over time, 2) group differences, 3) correlation with outcomes, 4) individual profiles along the array, and 5) independency from anatomical distance.

### Stability over time

We revealed an overall stability of the FI throughout the analysis period from the 3^rd^ month to the 1^st^ year post-surgery (M3-Y1). The confirmation of stability was important in two regards. First, it enabled us to combine ears that were recorded at different time points within the defined period. Second, it fits the assumption that SGN survival in humans is usually changing only slowly (as opposed to animal models) over the course of several years, and was most significantly determined by etiology (for review see Shepherd & Hardie, 2002).

### Group differences based on etiology and age

We confirmed that the average FI (i.e., mean value over all assessed CI contacts of an ear) is sensitive to demographic factors associated with reduced SGN survival. Most notably, the FI was elevated in the elderly and in children with confirmed cochlear hypoplasia.

The significantly higher FI in CI users with cochlear hypoplasia aligns with previous findings showing that the eCAP AGFs in paediatric patients with cochlear-nerve deficiency have shallower slopes and smaller amplitudes than children with normal cochlear-nerve anatomy (He et al., 2018). This further corresponds to histopathological observations of reduced SGN counts in this patient group (Nadol Jr, 1997).

In CI users with idiopathic etiologies, the mean FI was significantly elevated from 60 years onwards compared to younger CI users. Furthermore, we found a significant age-related increase for ears with a genetic cause of deafness (i.e., the only common etiology for pre- and post-lingual ears in the study population). These findings correspond to other studies, reporting age-related changes of eCAP parameters (e.g., Zamaninezhad et al., 2023) as well as temporal bone studies, describing age-related reduction in SGN counts (Kusunoki et al., 2004).

The variability in FI for different causes of deafness was high, likely reflecting the general variability in SGN loss. Correspondingly, age-normative SGN loss can range from 28% to 100% in patients with hereditary deafness (Teufert et al., 2006)

### Individual profiles along the CI array

We did not only confirm the usefulness of averaged FI for the group analyses, but also verified the site specificity of FI-values for individual contacts. We concentrated the analysis on FI patterns from ears with overall high FI values, as we propose that in these cases the maximal FI along the array were indicative of areas with substantially reduced SGN survival. Corresponding to the literature (DeVries et al., 2016; Pfingst et al., 2004; Wohlbauer et al., 2024; Zhou & Pfingst, 2014) we found individually distinct profiles along the array. Thereby, the maxima were most often located at the most basal contact. This corresponds to findings in temporal bone studies, describing aggravated SGN loss in basal/ high frequency areas of the cochlea (for review see Shepherd & Hardie, 2002).

Contrasting to the overall stability in FI, we found elevation with time for these cases, with high average FI-value. We propose that this may indicate that the rate of SGN degeneration is accelerated for ears which have low neural health already at/ shortly after implantation. This aspect should be analysed further with a higher sample size as it could help to manage outcome expectations with the CI.

### Correlation with speech perception

The most direct (i.e., still indirect) confirmation of the FI as neuronal marker in humans is a significant correlation with speech perception. In accordance with the animal model (Konerding et al., 2025), we used maximal FI-values per ear to assess correlation with severity of the neuronal damage.

The most common speech test in our data set was the Freiburger monosyllabic word test in quiet. The variance in FI explained about 12% of its variance. The useability of the monosyllabic word test for assessing neural markers has been debated: Although some studies described a positive correlation between total SGN count and CNC word recognition (e.g., Kamakura & Nadol Jr, 2016), a meta-analysis concluded that monosyllabic word scores were not correlated with SGN survival (Cheng & Svirsky, 2021). However, a recent study found that the variance in estimated number of fibres (as per eCAP recording) explained about 12% of variance in the monosyllabic speech test (Dong et al., 2023). We therefore conclude that our findings are within the expected range for a marker of SGN survival for this type of perceptual task (e.g., 9-12 % in Skidmore et al., 2023).

We described the strongest correlation between the FI and the speech-in-noise task. The variance in FI explained about 55% of the variance in HSM in noise scores (r^2^=0.545). A similarly high coefficient of determination has been described for a different eCAP measure, the channel-separation index (Scheperle & Abbas, 2015). However, this measure is time consuming (i.e., requiring recording with different combinations of masker and probe) and requires relative assessment of electrode pairs. Furthermore, the correlation was observed for speech perception using an experimental speech processor map, rather than the map used in everyday practice. Another approach, resulting in correlation coefficients of about 0.6, is to assess the deviation from the typical relation between eCAP threshold and EMD (Long et al., 2014). This approach is, however, dependent on the availability of post-operative CT scans of sufficient quality. The reported strength of correlation (r^2^=0.6) is likely reaching the upper limit of how much of the variance in speech understanding is explicable by peripheral factors. Thus, higher values have only been reported for within-subject designs, assessing differences between ears (e.g., Schvartz-Leyzac & Pfingst, 2018).

Commonly, studies that report significant correlations between speech-in-noise scores and electrophysiological markers report coefficients of determination of slightly above 0.3 (Dawson et al., 2025; Dong et al., 2023; Mesnildrey et al., 2020; Schvartz-Leyzac et al., 2025; Skidmore et al., 2022, 2023). This value corresponds well to the one we found for the maximal eCAP amplitude, verifying that our reduced data set for speech-correlation was a representative sample population. Therefore, we conclude that the strong correlation of the FI with speech-in-noise perception indicates a high validity of the FI as a neural health marker.

### Independence from distance to excitable structures

A recent study assessed a potential approximation of the FI: the input/output ratio at most comfortable level (MCL; Garcia & Carlyon, 2025). Whereas this FI_MCL_ could identify simulated cochlear dead regions, it was also significantly affected by EMD. As that study deviated from the original calculation of the FI, we assessed in our data set, to what extent the different eCAP parameters were influenced by EMD. Concordantly with the proposed rationale of the original FI, the FI was independent from the distance to the excitable structures (as estimated by EMD to the modiolar wall).

Corresponding to the literature (Degen et al., 2020; Lee et al., 2021) we found significant elevation in threshold with increasing distance to the modiolus and a significant effect of EMD on maximal eCAP amplitude. Hereby it was interesting to note that the maximal amplitude peaked at an EMD of around 1.4 mm. To the best of our knowledge, this relation has not been reported before.

### Methodical Considerations

We established the FI as the input/output ratio at saturation level (i.e., the inverse of, but not interchangeable with an efficiency index; see Appendix 2). The saturation amplitude (max amplitude) indicates the maximal excitable neural population and the respective charge (charge_max_) indicates the minimal charge necessary to activate them. Thereby, a high FI reflects the need to use high charge levels to even excite a small number of healthy neurons. As measurements have to be pre-terminated if stimulation levels are perceived too loud, saturation levels are not always reached. Therefore, recent publications approximated FI at MCL (Garcia & Carlyon, 2025; Wohlbauer & Arenberg, 2025). As discussed above, this FI_MCL_ is not optimal as a neural health marker. However, it is not necessary to record the saturation amplitude directly, as it is inferred from the sigmoidal function (formula 1). Due to the point reflection of the sigmoidal AGF function it can be reliably estimated already when the stimulation levels are high enough to reach the inflection point of the sigmoidal curve (Konerding et al., 2025). The inflection point in our subjects was on average 18 nC (SD: 4nC, 7-44nC) and, thus, reasonably low for eCAP recordings.

Due to the fact that we included a comparison to eCAP threshold, we restricted our analysis to those recordings, for which a threshold was detected. This introduced a quality criterion, as the automatic procedure in the recording software only provides a threshold, if the slope of the AGF function was reliably detected (Strahl, S. et al., 2018). Therefore, cases with lowest neural health were most likely excluded. Correspondingly, a study in a patient group with supposed high SGN degeneration reported elevated numbers of invalid eCAP recordings relative to matched controls (Moyaert et al., 2025). The assumption was supported by the distribution of speech understanding scores (Fig. 7), which was underrepresenting low performers (i.e., <40-70% correct depending on speech task). However, it has been shown that low eCAP amplitudes or even absence of eCAP responses is not significantly associated with poor speech recognition (Cooper et al. 2020). This is likely due to the fact that the lack of an evaluable eCAP amplitude (i.e., low signal-to-noise ratio) can also be due to low recording quality, resulting in increased noise. We are currently exploring extensions to the FI for recordings with insufficient signal-to-noise ratio to distinguish cases with very low neural health from those with bad recording quality.

We used the grand median over all averaged FI-values (i.e., mean FI for each ear) for dichotomization into supposedly healthy auditory nerves (<FI_GM_) and cochleae with reduced neural health (>FI_GM_). The limitation of this approach is that (other than in the animal model) we only included deafened ears and thus, cannot be certain that a sufficient number of healthy auditory nerves were included for a reliable median-split (for details see, Konerding et al., 2025). However, it has been described that even in the absence of hair cells, the number of SGNs can remain at 100% of age-normative cell numbers and about half of the ears remained within two standard-deviations of normal (Teufert et al., 2006). Still, we cannot rule out that the true lines between healthy and degenerated SGN may be below the indicated FI_GM_. However, as the hypoplasia group were all at or above the FI_GM_ (Fig. 5B) and the average FI was changing from below to above this value throughout aging (Fig. 4), we consider the FI_GM_ a helpful tool to identify ears and groups with substantially reduced SGN loss.

One limitation of this work is the retrospective nature of the study: Although all results indicate that the FI is a valuable marker of neural health, the testing remained indirect (i.e., no direct estimation of SGN counts was available). The next step is to assess to what extent the FI can be used as a marker to guide tailored programming strategies for individual CI users. As has been reviewed by Carlyon and Goehring (2021), individualized speech-processor programming in regions of supposed bad neural function (e.g., deactivating subsets of electrodes) has yielded mixed results, which may be due to the limitation of previously tested markers of neural health. If successful, ongoing prospective experiments will not only give direct validation of the FI as neural health marker, they might also provide the audiologist with a reliable criterion for electrode selection in clinical speech-processor programming (Sander et al., 2023).

## Conclusion

We conclude that the FI is a valuable marker for neural health, suited to inform about SGN loss, both at the group level, and for individual ears, and even for individual CI contacts. It is applicable for a wide variety of CI users, with different etiologies and demographic background, for both the paediatric and adult population (pre- and post-lingually deafened). The calculation of the FI is simple and requires only the standard AGF parameters assessable via clinical setups without any additional equipment or time-investment needed. We propose that average FI-values, in combination with the median splitting approach (FI_GM_), can assist in the identification of individual ears with special need for support during rehabilitation, and the FI patterns along the array may guide speech processor fitting procedures to optimize speech outcomes with a CI.

## Data Availability

All original data is stored in the database of the German Hearing Center. Pre-processed data, without information on the individual patients, can be shared upon request.

## Acknowledgments

We thank the clinical teams of the ENT department at MHH and the audiologists and therapists at the German Hearing Center for their work which was the basis for the data included in this study.

## Statements and declarations

### Ethical considerations

This publication contains data collected during the clinical routine and is approved by the ethics board of the Hannover Medical School (No. 1897-2013). The ethics board approval covers the evaluation of all data acquired during the clinical routine. The ethics committee waived the requirements for informed consent.

### Consent to participate

Not applicable.

### Consent for publication

Not applicable.

### Declaration of conflict of interest

CB, HB, OO, DS, SV, SS, JT, and CG are employed by MED-EL Medical Electronics GmbH with scientific roles only and no marketing or sales activities. All other authors report no conflict of interest.

## Funding

The work was supported by MHH-plus foundation. Besides the financial support, the founding agency had no involvement in the study.

