## Appendix 2 for "The Failure Index as marker of cochlear health in MED-EL CI users: Anatomy, demographics, and speech correlations"

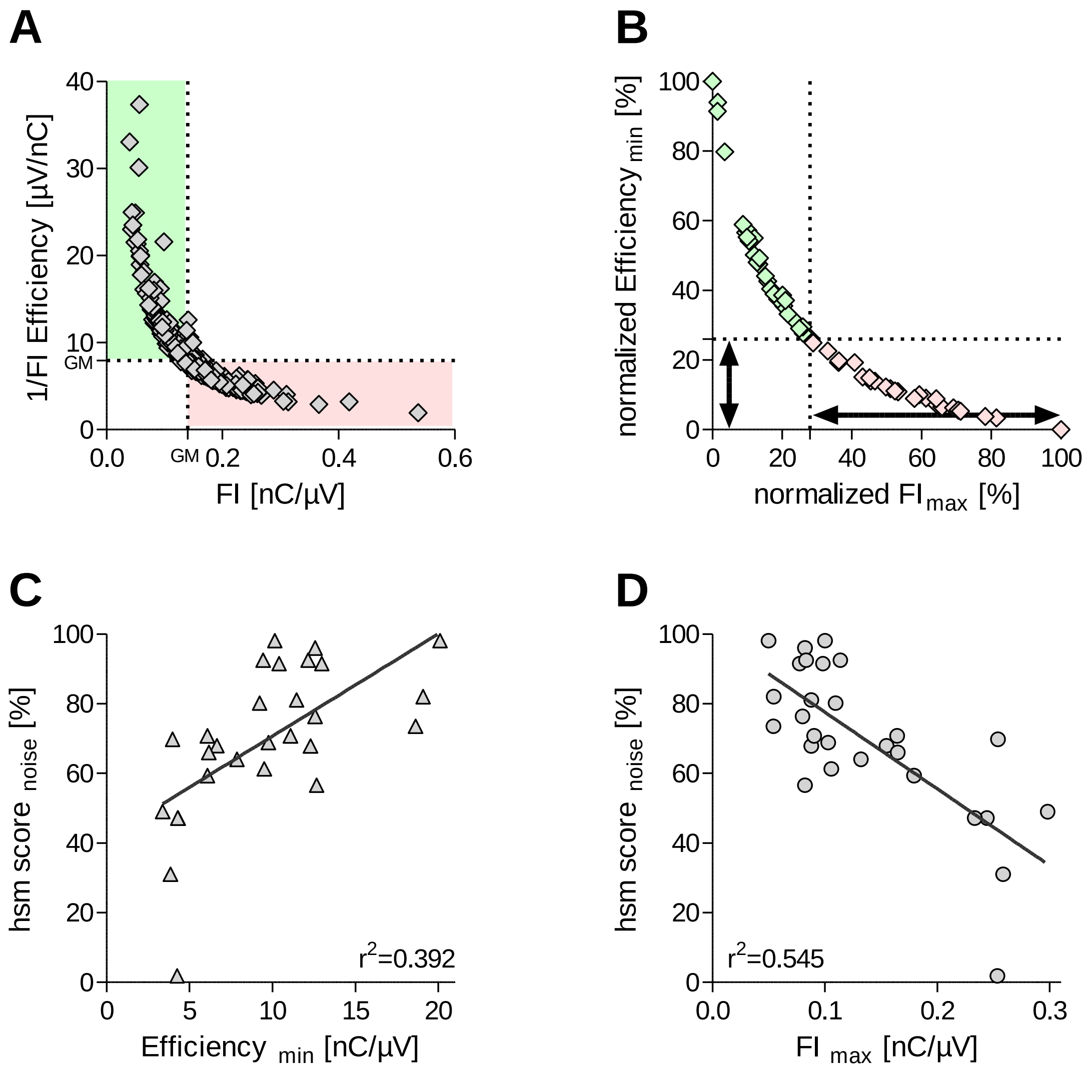


**Appendix 2 – Although the FI is the inverse of an Efficiency Index (1/FI), these two are not interchangeable.**

A) The mean FI and the mean Efficiency for each ear (N=278) do usually, but not in every case, lead to similar conclusions on presumed health ear (green shading) and ears with presumed reduced neural health (pink shading) when applying a split at the respective grand medians (GM). B) The min-max normalization of extreme values along the array (n=55) highlights the main difference between the two measures: The presumed affected ears (pink symbols) cover more than 70% along the FI dimension (vertical arrow), but less than 30% along the Efficiency dimension (horizontal arrow). Therefore, (C) the variance in Efficiency explains less than 40% of the variance in speech in noise perception, whereas, (D) the variance in FI explains nearly 55% of it (same n=28 ear in C&D).
