## Appendix 1 for "The Failure Index as marker of cochlear health in MED-EL CI users: Anatomy, demographics, and speech correlations"

| Patient | language | sex | ear | status | aetiology | steroid | disability | implant | age* |
|---|---|---|---|---|---|---|---|---|---|
| *1* | bilingual | F | R | pre | unknown | | | Flex 28 | <20 |
| | | | L | pre | unknown | | | Flex 28 | <20 |
| *2* | German | F | L | post | Menière's disease | | | Flex 24 | 60-69 |
| *3* | other | F | R | post | unknown | | | Flex 28 | 70-79 |
| *4* | German | M | R | pre | unknown | | | Flex 28 | <20 |
| | | | L | pre | unknown | | | Flex 28 | <20 |
| *5* | German | F | R | post | pertussis | | | Flex 28 | 60-69 |
| *6* | German | F | L | post | traumatic | y | | Flex 28 | 30-39 |
| *7* | German | M | R | post | unknown | | | Flex 28 | 60-69 |
| *8* | German | M | L | post | Cholesteatom/SHL | | | Flex 28 | 70-79 |
| *9* | German | M | L | post | unknown | | | Flex 24 | 80-91 |
| *10* | German | M | R | post | unknown | | | Flex 28 | 80-91 |
| *11* | bilingual | F | R | post | SHL | | | Flex 28 | 60-69 |
| *12* | German | M | R | post | unknown | | | Flex 24 | 70-79 |
| *13* | German | F | R | post | Wegener's | y | | Flex 28 | 70-79 |
| *14* | German | F | R | post | unknown | | | Flex 28 | <20 |
| ***15*** | **German** | **M** | **R** | **post** | **SHL** | **y** | | **Flex 28** | 20-29 |
| *16* | German | F | R | post | unknown | | | Flex 24 | 50-59 |
| *17* | German | M | L | post | unknown | | | Flex 28 | 80-91 |
| *18* | German | F | L | pre | genetic | | | Flex 28 | <20 |
| | | | R | pre | genetic | | | Flex 28 | <20 |
| *19* | German | F | L | post | SHL | | | Flex 24 | 20-29 |
| ***20*** | **German** | **M** | **L** | **post** | **unknown** | **y** | | **Flex 28** | 80-91 |
| *21* | German | M | R | post | acoustic neuroma | | | Flex 28 | 50-59 |
| *22* | German | F | R | post | unknown | | | Flex 28 | 60-69 |
| *23* | other | M | L | pre | unknown | y | | Flex 28 | 30-39 |
| *24* | German | M | R | post | unknown | | | Flex 28 | 60-69 |
| *25* | German | M | L | post | unknown | y | | Flex 28 | 80-91 |
| *26* | bilingual | M | L | pre | genetic | | | Flex 28 | <20 |
| *27* | German | F | R | post | otitis media | | | Flex 28 | <20 |
| *28* | other | F | R | post | unknown | | | Flex 24 | 20-29 |
| *29* | German | M | R | post | Menière's disease | y | | Flex 28 | 70-79 |
| *30* | German | M | L | post | SHL | | | Flex 28 | 30-39 |
| *31* | German | F | R | post | unknown | | | Flex 24 | 70-79 |
| *32* | German | F | R | pre | unknown | | | Flex 28 | <20 |
| *32* | | | L | pre | unknown | | | Flex 28 | <20 |
| *33* | other | M | R | post | unknown | y | | Flex 28 | 60-69 |
| *34* | German | F | L | post | unknown | | | Flex 28 | 50-59 |
| *35* | other | M | L | pre | unknown | | | Flex 28 | <20 |
| *36* | German | M | R | post | traumatic | | | Flex 28 | 70-79 |
| *37* | bilingual | F | L | post | SHL | y | | Flex 28 | 60-69 |
| *38* | German | F | R | post | Menière's disease | y | | Flex 28 | 40-49 |
| *39* | German | F | R | pre | Cytomegalovirus | | | Flex 28 | <20 |
| *40* | German | M | L | post | traumatic | | | Flex 28 | 50-59 |
| *41* | German | M | L | pre | hypoplasia | | | Flex 28 | <20 |
| *42* | German | F | R | post | SHL | | | Flex 28 | 40-49 |

| | | | | | | | | | |
|---|---|---|---|---|---|---|---|---|---|
| 43 | German | M | R | post | unknown | | | Flex 28 | 50-59 |
| 44 | German | M | R | post | genetic | | | Flex 28 | 50-59 |
| 45 | German | F | R | post | unknown | | | Flex 28 | 20-29 |
| **46** | **German** | **F** | **L** | **post** | **genetic** | **y** | | **Flex 28** | 60-69 |
| 47 | German | F | R | post | unknown | y | | Flex 28 | 70-79 |
| 48 | German | M | L | post | unknown | | y | Flex 24 | 50-59 |
| 49 | German | F | L | post | unknown | | y | Flex 28 | 80-91 |
| 50 | bilingual | M | L | pre | genetic | | | Flex 28 | <20 |
| | | | R | pre | genetic | | | Flex 28 | <20 |
| 51 | German | M | L | post | genetic | | | Flex 28 | 60-69 |
| 52 | other | F | R | pre | unknown | | | Flex 28 | <20 |
| 53 | German | M | R | post | unknown | | | Flexsoft | 70-79 |
| 54 | German | M | R | post | unknown | | | Flex 28 | 60-69 |
| 55 | other | F | R | post | unknown | | y | Flexsoft | 60-69 |
| 56 | German | M | R | post | Meningitis | | | Flex 28 | 60-69 |
| 57 | German | F | L | post | SHL | | | Flex 28 | 80-91 |
| 58 | German | M | R | pre | unknown | | | Flex 28 | 50-59 |
| 59 | German | F | R | post | SHL | | | Flex 28 | 60-69 |
| 60 | unknown | F | L | post | SHL | | | Flex 28 | 40-49 |
| 61 | German | F | R | pre | hypoplasia | | | Flex 28 | <20 |
| **62** | **German** | **F** | **L** | **post** | **unknown** | | | **Flex 28** | 80-91 |
| **63** | **German** | **F** | **R** | **post** | **SHL** | **y** | | **Flex 28** | 50-59 |
| **64** | **German** | **M** | **R** | **post** | **SHL** | **y** | | **Flexsoft** | 50-59 |
| **65** | **German** | **F** | **L** | **post** | **unknown** | **y** | | **Flex 28** | 70-79 |
| 66 | bilingual | F | R | post | Cogan-I-Syndrom | | | Flex 28 | 50-59 |
| 67 | German | F | L | post | SHL | | | Flex 28 | 80-91 |
| 68 | German | F | L | post | unknown | | y | Flex 28 | 50-59 |
| 69 | German | F | R | post | unknown | | | Flex 28 | 80-91 |
| 70 | German | F | L | post | unknown | | | Flex 28 | 50-59 |
| 71 | other | M | R | pre | unknown | | | Flex 28 | <20 |
| 72 | German | F | L | post | unknown | | y | Flex 28 | 80-91 |
| 73 | other | M | R | pre | unknown | | y | Flex 28 | <20 |
| | | | L | pre | unknown | | y | Flex 28 | <20 |
| **74** | **German** | **M** | **L** | **post** | **unknown** | **y** | | **Flex 28** | 50-59 |
| 75 | German | M | L | pre | unknown | | | Flex 28 | <20 |
| 76 | other | F | R | post | unknown | | | Flex 24 | <20 |
| 77 | German | F | L | pre | LVA-Syndrom | | | Flex 28 | <20 |
| 77 | | | R | pre | LVA-Syndrom | | | Flex 28 | <20 |
| 78 | German | F | R | pre | unknown | | | Flex 28 | <20 |
| 79 | German | M | L | post | unknown | | | Flex 28 | 80-91 |
| 80 | German | M | L | post | unknown | | | Flex 28 | 70-79 |
| 81 | German | M | R | post | unknown | | | Flex 28 | 70-79 |
| **82** | **German** | **F** | **L** | **post** | **unknown** | **y** | | **Flex 28** | 30-39 |
| | | | **R** | **post** | **unknown** | **y** | | **Flex 28** | 30-39 |
| 83 | German | F | L | post | unknown | | | Flex 28 | 60-69 |
| 84 | German | F | R | Post | unknown | | | Flex 28 | 60-69 |
| 85 | German | F | R | Post | unknown | | | Flex 28 | 50-59 |

| | | | | | | | | | |
|---|---|---|---|---|---|---|---|---|---|
| *86* | German | M | L | post | SHL | | | Flex 28 | 50-59 |
| ***87*** | **German** | **F** | **L** | **post** | **SHL** | | | **Flex 28** | 30-39 |
| ***88*** | **German** | **M** | **R** | **post** | **unknown** | **y** | | **Flex 28** | 80-91 |
| *89* | German | F | R | pre | unknown | | | Flexsoft | <20 |
| | | | L | pre | unknown | | | Flexsoft | <20 |
| *90* | German | M | L | pre | unknown | | | Flex 28 | <20 |
| *91* | German | F | L | post | SHL | | | Flex_28 | 30-39 |
| *92* | German | M | L | post | Menière's disease | | | Flex_28 | 60-69 |
| *93* | bilingual | F | R | pre | unknown | | | Flex 28 | <20 |
| *93* | | | L | pre | unknown | | | Flex 28 | <20 |
| ***94*** | **German** | **F** | **L** | **post** | **SHL** | | | **Flex_28** | 50-59 |
| *95* | German | M | R | Post | otitis media | | | Flex_26 | 70-79 |
| *96* | bilingual | M | L | post | traumatic | | y | Flex_28 | 50-59 |
| *97* | other | M | L | pre | hypoplasia | | | Flex 26 | <20 |
| *98* | other | M | R | pre | unknown | | | Flex 28 | <20 |
| | | | L | pre | unknown | | | Flex 28 | <20 |
| *99* | German | F | L | post | SHL | | | Flex 28 | 70-79 |
| *100* | German | M | L | post | unknown | | | Flex_28 | 70-79 |
| *101* | German | F | L | post | unknown | | | Flex_28 | 70-79 |
| *102* | German | F | L | post | unknown | y | | Flex 28 | 50-59 |
| ***103*** | **German** | **M** | **L** | **Post** | **unknown** | **y** | | **Flex 28** | 60-69 |
| *104* | German | F | L | post | unknown | | | Flex_28 | 40-49 |
| *105* | German | F | L | pre | unknown | | | Flexsoft | 40-49 |
| *106* | other | M | L | pre | unknown | | | Flexsoft | <20 |
| *106* | | | R | pre | unknown | | | Flexsoft | <20 |
| *107* | German | M | L | post | unknown | | | Flex_26 | 70-79 |
| *108* | German | M | L | post | SHL | | | Flexsoft | 50-59 |
| *109* | bilingual | M | L | post | unknown | | | Flex_28 | <20 |
| *110* | German | M | R | pre | unknown | | | Flex 28 | <20 |
| | | | L | pre | unknown | | | Flex 28 | <20 |
| *111* | German | F | L | post | unknown | | | Flex_28 | 80-91 |
| *112* | German | F | L | post | unknown | | y | Flex_28 | 50-59 |
| *113* | bilingual | M | R | pre | unknown | | | Flex 28 | <20 |
| *114* | German | M | L | post | SHL | | | Flex_28 | 50-59 |
| *115* | German | F | R | Post | SHL | | | Flex_28 | 60-69 |
| *116* | bilingual | F | L | post | unknown | | | Flex 28 | 60-69 |
| *117* | German | F | L | pre | hypoplasia | | | Flex 28 | <20 |
| *118* | German | F | R | pre | unknown | | | Flexsoft | <20 |
| *118* | | | L | pre | unknown | | | Flexsoft | <20 |
| *119* | bilingual | F | R | pre | unknown | | | Flex 26 | <20 |
| *120* | bilingual | F | R | pre | unknown | | | Flex 28 | <20 |
| | | | L | pre | unknown | | | Flex 28 | <20 |
| *121* | other | F | R | pre | unknown | | | Flex 28 | <20 |
| *122* | German | F | R | post | Menière's disease | | | Flexsoft | 70-79 |
| *123* | German | F | R | post | unknown | | | Flex_28 | 80-91 |
| *124* | German | M | L | pre | unknown | | | Flex_28 | 50-59 |
| *125* | German | M | L | post | unknown | y | | Flex_28 | 50-59 |

| | | | | | | | | | |
|---|---|---|---|---|---|---|---|---|---|
| *126* | German | M | R | Post | unknown | | | Flex 24 | 60-69 |
| *127* | German | F | L | post | unknown | | | Flexsoft | 70-79 |
| *128* | German | F | R | pre | hypoplasia | | | Flex 28 | <20 |
| ***129*** | **German** | **M** | **R** | **Post** | **ototoxicity** | | | **Flex_28** | 60-69 |
| *130* | German | F | L | pre | unknown | | y | Flexsoft | <20 |
| *131* | German | M | R | post | unknown | | | Flexsoft | 70-79 |
| *132* | German | F | L | Post | unknown | | | Flex_28 | 60-69 |
| *133* | other | F | R | pre | unknown | | y | Flexsoft | <20 |
| | | | L | pre | unknown | | y | Flexsoft | <20 |
| *134* | German | M | R | post | acoustic neuroma | | | Flex_28 | 70-79 |
| *135* | German | M | L | post | cortical infarct | | | Flex_28 | 70-79 |
| *136* | German | F | R | post | SHL | | | Flex_28 | 50-59 |
| *137* | German | M | R | Post | tumor | | | Flexsoft | 60-69 |
| *138* | German | M | L | pre | unknown | | | Flex 28 | <20 |
| *139* | German | F | L | post | unknown | | | Flex_28 | 60-69 |
| *140* | German | F | L | post | Menière's disease | | y | Flex_28 | 50-59 |
| *141* | N/A | M | R | pre | unknown | | | Flex_28 | <20 |
| *142* | German | F | R | pre | hypoplasia | | | Flex_28 | <20 |
| *143* | German | F | L | post | unknown | | | Flex_28 | 60-69 |
| *144* | German | M | L | post | unknown | | | Flex_28 | 80-91 |
| *145* | bilingual | F | R | pre | Cytomegalovirus | | | Flex_28 | <20 |
| | | | L | pre | Cytomegalovirus | | | Flex_28 | <20 |
| *146* | German | F | L | post | unknown | | | Flex_28 | 40-49 |
| *147* | German | F | L | pre | hypoplasia | | | Flex_28 | <20 |
| *148* | bilingual | F | L | post | chronic mastoiditis | | | Flex_28 | 50-59 |
| *149* | German | M | L | post | Waardenburg syndrome | | | Flex_28 | 40-49 |
| *150* | other | F | L | post | SHL | | | Flex_28 | 30-39 |
| *151* | German | F | L | post | SHL | | | Flex_26 | 70-79 |
| ***152*** | **German** | **M** | **L** | **post** | **SHL** | | | **Flex 28** | 70-79 |
| *153* | bilingual | F | L | post | unknown | | | Flex_28 | 50-59 |
| *154* | German | F | R | post | schwannoma | | y | Flex_28 | 60-69 |
| *155* | German | M | L | pre | unknown | | | Flex_28 | <20 |
| *156* | German | F | L | post | SHL | | | Flex_28 | 60-69 |
| ***157*** | **German** | **F** | **L** | **post** | **unknown** | | | **Flex_28** | 50-59 |
| *158* | other | F | L | post | Menière's disease | | | Flex_28 | 50-59 |
| *159* | German | M | L | post | SHL | | y | Flex_28 | 60-69 |
| ***160*** | **German** | **F** | **L** | **post** | **Menière's disease** | | | **Flex_28** | 50-59 |
| *161* | German | M | R | post | traumatic | | | Flex_28 | 40-49 |
| ***162*** | **German** | **F** | **R** | **post** | **unknown** | **Y** | | **Flex 28** | 70-79 |
| *163* | German | F | R | post | unknown | | | Flex_28 | 40-49 |
| *164* | German | F | L | post | unknown | | | Flex_28 | 60-69 |
| ***165*** | **German** | **M** | **R** | **post** | **unknown** | | | **Flex_28** | 20-29 |
| *166* | German | F | L | post | unknown | | y | Flex_28 | 80-91 |
| *167* | German | M | L | post | unknown | | y | Flex_28 | 60-69 |
| ***168*** | **German** | **M** | **R** | **post** | **SHL** | | | **Flex_28** | 60-69 |

| | | | | | | | | | |
|---|---|---|---|---|---|---|---|---|---|
| *169* | German | F | L | post | genetic | | | Flex_28 | 30-39 |
| ***170*** | **German** | **F** | **L** | **post** | **SHL** | | | **Flex_28** | 50-59 |
| *171* | German | F | R | post | SHL | | y | Flex_28 | 50-59 |
| *172* | German | M | R | post | ototoxicity/ Menière's | | | Flexsoft | 70-79 |
| *173* | German | F | L | pre | genetic | | | Flex_28 | <20 |
| | | | R | pre | genetic | | | Flex_28 | <20 |
| *174* | other | F | L | post | SHL | | | Flex_28 | 60-69 |
| *175* | German | F | R | post | unknown | | | Flex_28 | 60-69 |
| ***176*** | **German** | **F** | **R** | **post** | **unknown** | **Y** | | **Flex 28** | 30-39 |
| *177* | German | F | R | post | unknown | | | Flex_28 | 70-79 |
| *178* | bilingual | M | R | post | unknown | | | Flex_28 | 50-59 |
| ***179*** | **German** | **F** | **R** | **post** | **unknown** | **Y** | | **Flex 28** | 50-59 |
| *180* | German | F | L | post | SHL | | y | Flex_28 | 50-59 |
| *181* | German | F | R | post | unknown | | | Flex_28 | 50-59 |
| *182* | German | M | R | pre | unknown | | | Flex_28 | <20 |
| | | | L | pre | unknown | | | Flex_28 | <20 |
| *183* | other | F | L | post | Scarlatina | | | Flex_28 | 30-39 |
| ***184*** | **German** | **F** | **L** | **post** | **unknown** | | | **Flex_28** | 50-59 |
| ***185*** | **German** | **M** | **R** | **post** | **Sepsis** | **Y** | | **Flex 28** | 20-29 |
| *186* | other | M | L | pre | genetic | | y | Flex_28 | 20-29 |
| *187* | bilingual | M | R | post | unknown | | | Flex_28 | 50-59 |
| ***188*** | **German** | **F** | **L** | **post** | **Menière's disease** | | | **Flex_28** | 50-59 |
| *189* | other | F | R | post | unknown | | | Flex_28 | 30-39 |
| *190* | German | F | L | post | unknown | | | Flex_28 | <20 |
| ***191*** | **German** | **F** | **R** | **post** | **unknown** | | | **Flex_28** | 40-49 |
| *192* | German | M | R | post | unknown | | | Flex_28 | 40-49 |
| ***193*** | **German** | **M** | **R** | **post** | **SHL** | | | **Flex_28** | 70-79 |
| ***194*** | **German** | **M** | **R** | **post** | **unknown** | | | **Flex_28** | 80-91 |
| *195* | other | M | L | post | unknown | | | Flex_28 | 40-49 |
| *196* | German | F | R | pre | unknown | | | Flex 28 | <20 |
| | | | L | pre | unknown | | | Flex 28 | <20 |
| *197* | German | F | L | post | unknown | | | Flex_28 | 20-29 |
| *198* | German | M | L | post | SHL | | | Flex_28 | 50-59 |
| *199* | German | F | L | pre | unknown | | | Flex_28 | <20 |
| | | | R | pre | unknown | | | Flex_28 | <20 |
| *200* | other | F | L | post | unknown | | | Flex_28 | 70-79 |
| ***201*** | **German** | **M** | **L** | **post** | **unknown** | | | **Flex_28** | 80-91 |
| *202* | German | F | L | post | SHL | | | Flex_28 | 60-69 |
| *203* | German | F | R | post | Menière's disease | y | | Flex_28 | 30-39 |
| *204* | German | F | R | post | unknown | | | Flex_28 | 60-69 |
| *205* | bilingual | M | L | pre | unknown | | | Flex_28 | <20 |
| | | | R | pre | unknown | | | Flex_28 | <20 |
| *206* | bilingual | F | R | pre | unknown | | | Flex_28 | <20 |
| | | | L | pre | unknown | | | Flex_28 | <20 |
| ***207*** | **German** | **M** | **R** | **post** | **unknown** | | | **Flex_28** | 70-79 |
| *208* | German | M | L | post | unknown | | y | Flex_28 | 80-91 |

| | | | | | | | |
|---|---|---|---|---|---|---|---|
| *209* | bilingual | F | L | post | SHL | Flex_28 | 60-69 |
| ***210*** | **German** | **F** | **L** | **post** | **SHL** | **Flex_28** | 70-79 |
| *211* | German | M | L | post | unknown | Flex_28 | 80-91 |
| *212* | bilingual | M | R | post | traumatic | Flex_28 | 70-79 |
| *213* | German | F | L | post | unknown | Flex_28 | 50-59 |
| ***214*** | **German** | **M** | **R** | **post** | **unknown** | **Flex_28** | 60-69 |
| *215* | other | F | L | pre | unknown | Flexsoft | <20 |
| | | | R | pre | unknown | Flexsoft | <20 |
| *216* | German | F | L | post | unknown | Flex_28 | 70-79 |
| *217* | German | M | R | post | SHL | Flex_28 | 70-79 |
| *218* | German | M | R | post | unknown | Flex_28 | 60-69 |
| *219* | other | F | L | pre | unknown | Flex_28 | <20 |
| *220* | German | M | R | post | SHL | Flex_28 | 70-79 |
| *221* | German | M | L | post | SHL | Flex_28 | 50-59 |
| *222* | German | M | R | post | unknown | Flex_28 | 60-69 |
| ***223*** | **German** | **M** | **L** | **post** | **SHL** | **Flex_28** | 70-79 |
| ***224*** | **German** | **M** | **R** | **post** | **tumor** | **Flex_28** | 60-69 |
| *225* | German | F | L | pre | unknown | Flex_28 | <20 |
| *226* | German | M | R | post | unknown | Flex_28 | 70-79 |
| *227* | other | F | L | post | unknown | Flex_28 | 40-49 |
| *228* | German | M | R | post | SHL | Flex_28 | 80-91 |
| *229* | German | F | L | post | unknown | Flex_28 | 70-79 |
| *230* | bilingual | M | R | post | Labyrinthitis | Flex_28 | <20 |
| | | | L | post | Labyrinthitis | Flex_28 | <20 |
| ***231*** | **German** | **F** | **L** | **post** | **SHL** | **Flex_28** | 30-39 |
| *232* | bilingual | M | L | post | SHL | Flex_28 | 50-59 |
| *233* | German | F | R | post | unknown | Flex_26 | 60-69 |
| *234* | bilingual | M | R | pre | Cytomegalovirus | Flex_28 | <20 |
| ***235*** | **German** | **M** | **L** | **post** | **SHL** | **Flex_28** | 30-39 |
| ***236*** | **German** | **M** | **L** | **post** | **Menière's disease** | **Flex_28** | 70-79 |
| *237* | German | M | R | pre | unknown | Flex_28 | <20 |
| *238* | German | M | L | post | genetic | Flex_26 | 70-79 |
| ***239*** | **German** | **M** | **L** | **post** | **genetic** | **Flex_28** | 60-69 |
| *240* | German | F | L | post | SHL | Flex_28 | 60-69 |
| *241* | German | F | L | post | unknown | Flex_28 | 50-59 |
| ***242*** | **German** | **F** | **L** | **post** | **unknown** | **Flex_28** | 20-29 |
| *243* | German | F | L | post | unknown | Flex_28 | 30-39 |
| *244* | bilingual | F | R | pre | unknown | Flex_28 | <20 |
| *245* | German | F | L | pre | Dysplasia/SHL | Flex_26 | 20-29 |
| *246* | other | F | R | post | Menière's disease | Flex_28 | 70-79 |
| *247* | other | F | L | post | unknown | Flex_28 | 60-69 |
| *248* | German | M | L | post | unknown | Flex_28 | 50-59 |
| *249* | German | F | L | post | unknown | Flex_28 | 70-79 |
| *250* | German | M | R | post | unknown | Flex_28 | 60-69 |
| *251* | German | M | R | post | unknown | Flex_28 | 60-69 |
| *252* | other | F | L | post | unknown | Flex_28 | 60-69 |
| *253* | other | M | R | post | unknown | Flex_28 | <20 |

* age-range at implantation
